# Preventive Effect of *Diospyros kaki* on Intraocular Pressure and Dry Eye: A Randomized, Double-Blind, Placebo-Controlled Clinical Trial

**DOI:** 10.1101/2025.07.21.25331959

**Authors:** Hyung Bin Hwang, Wook-Bin Lee, Kyung-A Kim, Hong Ryul Ahn, Tae Kyeom Kang, Jong Jin Lee, Min Kyoung Kim, Kui Dong Kang, Sang Hoon Jung

**Affiliations:** Department of Ophthalmology, Incheon St. Mary’s Hospital, College of Medicine, The Catholic University of Korea, Incheon, Korea; Center for Natural Product Efficacy Optimization, Korea Institute of Science and Technology, Gangneung 25451, Republic of Korea; Department of Natural Product Applied Science, University of Science and Technology (UST), Daejeon, 34113, Republic of Korea; Division of Medical Oncology, Yonsei Cancer Center, Department of Internal Medicine, Yonsei University College of Medicine, Republic of Korea; Whanin Pharm Co., Ltd., Seoul, Korea; Oxford Eye Center, Incheon, Korea

**Keywords:** Dry eye disease, Intraocular pressure, Diospyros Kaki, Functional foods, Clinical trial

## Abstract

**Background:** Dry eye disease (DED) is a common chronic ocular disorder associated with ocular discomfort and tear film damage. Traditional medicinal uses of *Diospyros kaki* leaves have been well-documented for their potent radical-scavenging, antioxidant, and immune-modulatory properties. Emerging evidence suggests that *D. kaki* leaves can improve various ocular conditions, including dry eye disease and elevated intraocular pressure (IOP), making it a potential therapeutic option for managing these conditions.

**Methods:** A randomized controlled trial was conducted involving one hundred patients diagnosed with mild DED. The participants were divided into placebo and treatment groups, receiving maltodextrin and EEDK (600 mg/day). Clinical efficacy, clinical indices, and adverse reactions were compared between the two groups.

**Results:** Administration of EEDK significantly improved tear film stability and corneal staining scores in patients with DED. IOP significantly decreased after EEDK administration. The EEDK group exhibited significant improvements in tear film stability compared with the placebo group at 4, 8, and 12 weeks. Consistent reductions in IOP and corneal staining scores were observed throughout the 12-week study period in the EEDK group. Moreover, EEDK intake effectively enhanced tear production as evidenced by the Schirmer test. No significant adverse events were observed.

**Conclusions:** The oral administration of EEDK demonstrated significant therapeutic effects in improving dry eye symptoms without notable adverse events. The results of this study suggest that EEDK could be considered an ideal therapeutic option for treating patients with dry eye disease.

**Trial registration:** The trial is registered with the Korean Clinical Trial Registry, KCT0010532

**Highlights:**

- Ethanol extract of *Diospyros kaki* (EEDK) improves dry eye symptoms and tear film stability.
- EEDK significantly reduces intraocular pressure in patients with dry eye disease.
- Randomized, double-blind, placebo-controlled clinical trial conducted over 12 weeks.
- No significant adverse events observed, indicating EEDK is well-tolerated.

**Graphical abstract:** 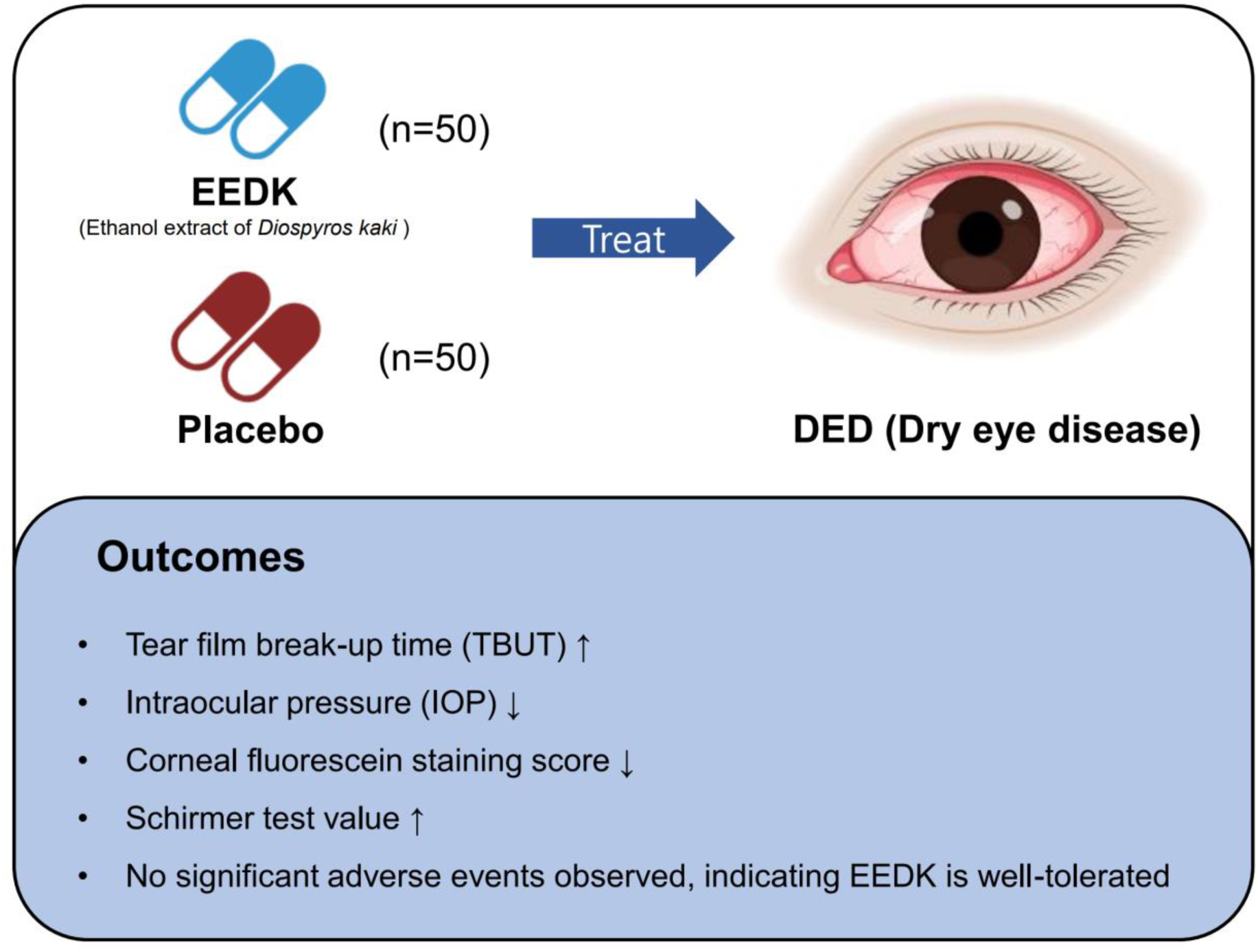

## 1. Introduction

Dry eye disease (DED) is a prevalent condition characterized by an abnormal tear film that leads to ocular surface damage and discomfort. It can be caused by tear deficiency or excessive tear evaporation, which affects various components involved in tear secretion and tear film formation [1, 2]. DED is associated with various factors such as aging, autoimmune diseases (e.g., rheumatoid arthritis, Sjogren’s syndrome, lupus, and diabetes), tear gland dysfunction, inflammation, conjunctivitis, thyroid disease, and medication use. Patients with DED frequently experience reduced visual acuity, impaired contrast sensitivity, and decreased quality of [3, 4].

Early detection and adequate treatment are crucial for managing DED [5]. Current therapeutic approaches focus on symptomatic relief by using artificial tears, anti-inflammatory agents, and essential fatty acid supplementation [6]. However, these treatments only provide temporary relief without addressing the underlying tear film dysfunction, resulting in unsatisfactory outcomes [7]. Moreover, preservatives used in eye drops can cause secondary damage to the ocular surface [8]. Therefore, it is necessary to investigate alternative treatment options to enhance the tear film function and reduce DED symptoms.

Functional diets have received increasing attention as potential treatments in this context. Functional foods consumed as daily dietary supplements contain micronutrients known as functional substances [9]. Scientifically validated clinical trials are essential to evaluate the efficacy and safety of functional foods containing these essential constituents for the treatment of DED.

One functional substance of interest is the ethanol extract of *Diospyros kaki* Thunberg (Ebenaceae), commonly known as persimmon. *D. kaki* leaves have been extensively studied and found to contain bioactive compounds, such as flavonoids, polyphenols, organic acids, and vitamins [10–13]. These compounds exhibit potent radical scavenging and antioxidant properties. Emerging evidence suggests that the medicinal effect of *D. kaki* leaves includes improving hemostasis, diuresis, constipation, and hypertension [13–16]. Animal experiments using *D. kaki* have demonstrated beneficial effects, such as optic nerve protection, angiogenesis inhibition, photoreceptor cell viability preservation, retinal thickness reduction, corneal protection, increased tear production, and decreased intraocular pressure [14, 17]. Our previous research showed promising results for the ethanol extract of *D. kaki* (EEDK) in improving dry eye disease in animal models.

This study aimed to evaluate the efficacy and safety of orally administered EEDK in patients with dry eye disease. By investigating the effects of EEDK in the human population, this study aimed to contribute to our understanding of its potential as a therapeutic option for managing dry eye disease.

## 2. Materials and Methods

### 2.1. Study Design

This prospective, randomized, double-blind clinical trial was conducted at Incheon St. Mary’s Hospital, Korea, from November 2019 to November 2020. A total of 100 patients diagnosed with mild-to-moderate dry eye disease (DED) were enrolled and randomly assigned to one of two groups: the test group received 600 mg of EEDK, and the control group received a placebo. Both the EEDK and placebo were provided by Whanin Pharm Co., Ltd. Participants were instructed to take two capsules of their assigned product twice daily for 12 weeks.

Clinical evaluations were conducted across four scheduled visits: baseline (week 0), week 4, week 8, and week 12. The primary outcomes included visual acuity, intraocular pressure (IOP), central corneal thickness, tear film breakup time (TBUT), Schirmer II test, noninvasive tear film breakup time (NIBUT), fluorescein corneal staining (assessed using the National Eye Institute grid), and ocular surface disease index (OSDI). Secondary outcomes, including blood biochemical and urine analyses, were evaluated at week 12. The trial was registered with the Clinical Research Information Service (CRIS) under registration number KCT0010532 (https://cris.nih.go.kr/cris/search/detailSearch.do?seq=29822).

### 2.2. Ethics and good clinical practice (GCP)

The study was conducted in accordance with the principles of the Declaration of Helsinki and Good Clinical Practice (GCP) guidelines. The study protocol was reviewed and approved by the Institutional Review Board (IRB) of The Catholic University of Korea (approval number: XC19HODE0043). All participants were recruited from a hospital-maintained database and provided written informed consent prior to study enrollment.

### 2.3. Study Patients

Patients meeting the following criteria were eligible for participation: (1) Age ≥19 years; (2) Diagnosed with mild to moderate dry eye and asymptomatic elevated intraocular pressure; (3) Intraocular pressure ≥21 mmHg in one or more eyes; (4) Cup/Disc (C/D) ratio < 0.5; (5) Tear break-up time (TBUT) < 10 s; and (6) Provided written informed consent prior to the start of the study.

Patients meeting any of the following criteria were excluded from the study: (1) Receiving treatment for eye diseases such as open-angle glaucoma (POAG), retinal disease, macular degeneration; (2) Glaucomatous optic nerve changes and visual field defects; (3) History of eye-related surgery (excluding cataract surgery) or eye-related disease (excluding dry eye syndrome) within 3 months prior to screening; (4) Risk factors for vision loss, except for diabetes and thyroid disease; (5) Systolic blood pressure of ≥ 160 mmHg or diastolic blood pressure of ≥ 100 mmHg (measured after 10 minutes of rest) or fasting blood sugar of ≥ 126 mg/dL at Visit 1 or Visit 2; (6) Use of drugs to regulate intraocular pressure within 3 months prior to screening; (7) Use of drugs known to affect intraocular pressure (e.g., steroids, airway dilators, statins, obesity drugs such as Topiramate, Phendimetrazine Tartrate, Mazindol, etc.); (8) Use of drugs known to affect dry eye syndrome (e.g., autoimmune disease treatments, etc.); (9) Use of contact lenses within 2 weeks prior to screening; (10) Consumption of health functional food or vitamin-containing health functional food that may affect eye health within 1 week prior to screening; (11) Heart disease requiring medication, surgery, or radiation therapy; (12) Sensitivity or allergy to the food used in the study; (13) Pregnant or planning to become pregnant during the study, or lactating; and (14) Participation in another clinical trial within 1 month prior to screening or plans to participate during the trial period.

### 2.4. Preparation of the drug and the placebo

The test product was an opaque, rigid capsule containing EEDK, whereas the placebo capsule was opaque and contained maltodextrin. Both the EEDK and placebo capsules were manufactured in a Good Manufacturing Practice pilot plant (Daepyung Co., LTD.). Stringent quality assurance measures were implemented during the manufacturing process to prevent contamination by microorganisms, heavy metals, and pesticide residues. Various measures were taken to ensure double-blinding. The production, packaging, and labeling of the test products and placebos used in this clinical trial were closely controlled to maintain consistency. Additionally, the allocation details, consisting of unique codes for each group, were kept in a sealed envelope in the custody of the designated clinical trial personnel. These codes were not disclosed until the completion of the clinical trial. The test capsules were provided by the clinical trial manager based on random codes assigned to each selected participant. To maintain blinding, spare capsules were available in case of defects or damage encountered during the trial.

### 2.5. Measurement of intraocular pressure

Intraocular pressure (IOP) was measured using slit-lamp-mounted Goldmann tonometry following administration of topical anesthesia with oxybuprocaine hydrochloride and sodium fluorescein.

### 2.6. Tear film break-up time and non-invasive tear film break-up time

A trained operator blinded to the treatment conditions carefully examined the ocular surface of both eyes of all participants to assess TBUT. To determine NIBUT at specific time intervals, video corneal topography (Idra, Essilor Ltd., Charenton-le-Pont, France) was used according to the manufacturer’s protocol. Participants were instructed to maintain a steady gaze, blink twice, and refrain from blinking for as long as possible. Upon opening the eye, data collection commenced, and the NIBUT was automatically recorded. Three consecutive measurements were obtained and the average of these measurements was used.

### 2.7. Corneal fluorescein staining

Corneal fluorescein staining was evaluated immediately after TBUT assessment. The evaluation was performed between 1 and 4 min after fluorescein instillation to prevent the dye from diffusing into the corneal stroma, which could obscure the boundary of any staining defects. The eyes were examined using a slit lamp at 16 × magnification, a yellow barrier filter, and cobalt blue illumination. Staining was graded according to the Oxford scale [18].

### 2.8. Schirmer II test

Schirmer’s test was used to assess tear production. Prior to administration of the anesthetic drops, Schirmer test strips were carefully positioned in the lower conjunctival sac at the intersection of the lateral and middle thirds to ensure no contact with the cornea. After a 5- minute interval, the length of the wet portion of the strips was measured in millimeters. The entire procedure was performed consistently by the same individual, at the same time and location. All patients were seated in a comfortable position with their eyes closed, and the lower cul-de-sac was gently dried using a cotton swab before strip placement.

### 2.9. Meibomian gland secretion (Meiboscore)

The meibomian gland score (Meiboscore) was used to quantify the scoring method established by Arita et al. [19] for the evaluation of meibomian gland atrophy. The Meiboscore was determined by evaluating changes in the meibomian glands of the upper and lower eyelids of each eye and adding these scores. A grade of 0 indicated no loss of meibomian glands, grade 1 represented an area of lost meibomian glands of less than 1/3 of the total area, and grade 2 indicated complete loss of meibomian glands. In cases where the area of loss was between 1/3 and 2/3, a grade of 3 was assigned. Once stability was attained, all measurements were performed by the same examiner using the same equipment at consistent time intervals.

### 2.10. Ocular surface disease index (OSDI)

The OSDI questionnaire was used to assess DED symptoms. The OSDI was developed by Allergan Inc. Outcome Research Group (Irvine, CA, USA). This questionnaire was specifically designed for individuals with DED and aimed to evaluate the frequency of specific symptoms and their impact on daily activities related to vision [20].

### 2.11. Evaluation of improvement by investigators and trial subjects

During visits 2 (week 4), 3 (week 8), and 4 (week 12), either the investigators or the trial subjects themselves assessed the degree of improvement in comparison to the baseline (before intake). A score of 1 indicated significant improvement in overall symptoms, a score of 2 indicated overall symptom improvement, a score of 3 indicated no change from baseline, a score of 4 indicated overall symptom worsening, and a score of 5 represented significant worsening in overall symptoms.

### 2.12. Adverse events

One hundred participants (50 in the test group and 50 in the control group) were included in the safety evaluation. The analysis included participants who were randomly assigned and had safety-related information collected after receiving at least one dose of the drug during the clinical trial. Adverse events including renal function damage, nausea, emesis, diarrhea, dizziness, and headache were assessed. Blood biochemistry and urine tests were also performed.

The incidences of adverse events reported during the trial period were also calculated. Before presenting the significance probability value, the ratio for the presence or absence of adverse reactions in each group was calculated, and the chi-square test (*χ*^2^ test) or Fisher’s exact test for independence was performed. Furthermore, the number of adverse reactions was tabulated, and an independence test was performed to evaluate the relevance of these cases to the health functional foods used in the clinical trial. The resulting significance probability values are reported.

### 2.13. Statistical analysis

All statistical analyses, including demographic characteristics, efficacy measurements, and safety evaluations, were conducted using one-sided tests with a significance level of 0.05. The reported *p*-values were presented up to four decimal places, and p < 0.05 was deemed statistically significant.

## 3. Results

### 3.1. Baseline Characteristics of Participants

A total of 100 patients diagnosed with DED met the inclusion criteria and were randomly assigned to either the EEDK treatment group (n=50) or the placebo group (n=50) prior to study initiation. Randomization was performed using a computer-generated randomization list. Among the participants, 11 who received at least one dose of treatment did not complete the trial and were subsequently excluded from the per-protocol set analysis (**Figure 1**). This left 89 participants (43 in the test group and 46 in the control group) for the per-protocol analysis, which was sufficient for statistical analysis. None of the participants had moderate or severe symptoms and there were no statistically significant differences between the treatment and placebo groups. No deaths or severe adverse events related to treatment were reported in any patient throughout the duration of the study.

**Figure 1.**
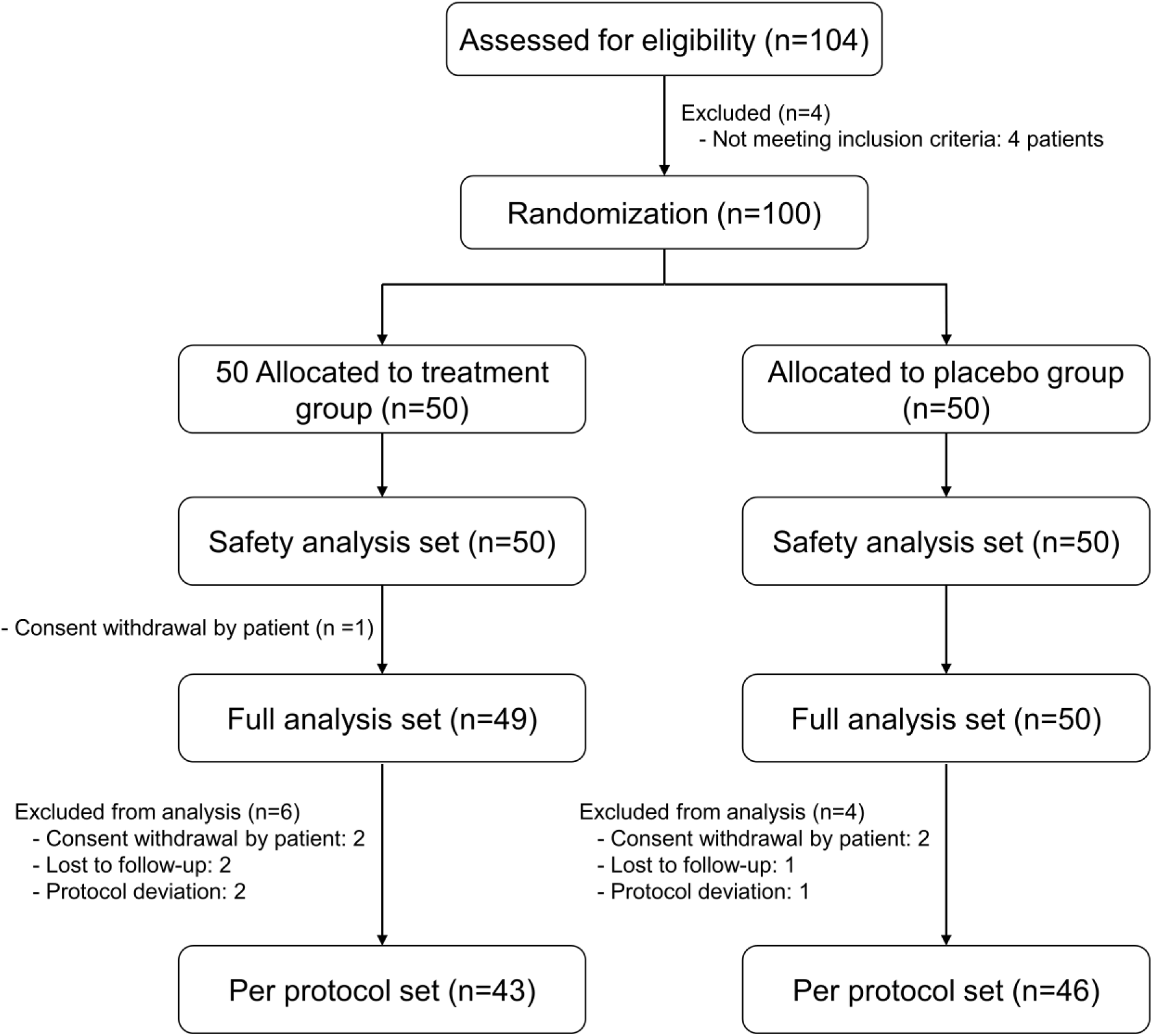
Flowchart of randomization and follow-up for the study (CONSORT flow diagram)

Before conducting any tests, demographic information was compared between the EEDK and placebo groups to identify any potential differences. **Table 1** summarizes the participants’ demographic information and other relevant characteristics before receiving the initial dose in the clinical trial. The EEDK group consisted of 11 men (25.58%) and 32 women (74.42%), whereas the placebo group consisted of 15 men (32.61%) and 31 women (67.39%). The mean age in the EEDK group was 56.63 ± 13.40 years, and in the placebo group, 58.17 ± 10.50 years (p = 0.7331). No statistically significant differences in these characteristics were observed between the two groups.

**Table 1.** Baseline patient characteristics.

| Characteristics | Experimental group (n=43) | Control group (n=46) | *p*-Value |
| --- | --- | --- | --- |
| Gender (male/female, n) | 11/32 | 15/31 | 0.4663 |
| Mean Age (y) | 56.63 ± 13.40 | 58.17 ± 10.50 | 0.7331 |
| Visual acuity | 0.81 ± 0.14 | 0.77 ± 0.17 | 0.1477 |
| Central Corneal Thickness (µm) | 547.86 ± 34.82 | 557.89 ± 35.85 | 0.1842 |
| TBUT | 4.02 ± 1.08 | 3.78 ± 0.99 | 0.2500 |
| IOP | 21.62 ± 0.43 | 22.16 ± 2.03 | 0.1490 |
| Schirmer test score (mm) | 8.00 ± 3.41 | 9.04 ± 3.75 | 0.1935 |
| NIBUT | 4.58 ± 0.71 | 4.55 ± 0.63 | 0.7300 |
| MGS | 1.56 ± 0.67 | 1.41 ± 0.62 | 0.2665 |
| Corneal fluorescein stain score | 3.77 ± 0.92 | 3.61 ± 0.77 | 0.3714 |
| OSDI | 27.69 ± 22.24 | 21.91 ± 20.04 | 0.1871 |

TBUT, tear film breakup time; IOP: Intraocular pressure; NIBUT, noninvasive tear film breakup time; MGS: Meibomian gland secretion; OSDI: Ocular surface disease index. There were no significant differences between the two groups at baseline.

### 3.2. The ethanol extract of *D. kaki* improved dry eye disease

**Figure 2** shows the changes in TBUT values at weeks 4, 8, and 12 after receiving EEDK and placebo. The treatment group that received EEDK exhibited improved tear film stability. At 4, 8, and 12 weeks after intake, there were significant differences in TBUT improvement between the EEDK and placebo groups (week 4, p = 0.0018; week 8, p = 0.0026; and week 12, p < 0.0001; **Table 2** and **Figure 2**). Compared with the baseline, TBUT in the EEDK group increased significantly by 1.49 ± 1.16 seconds (week 4), 1.51 ± 1.40 seconds (week 8), and 1.76 ± 1.17 seconds (week 12) (**Table 2**). In contrast, the placebo group showed only slight fluctuations in TBUT scores, which did not exhibit significant changes compared with baseline. The TBUT scores in the control group increased by 0.61 ± 1.41 seconds (week 4), 0.63 ± 1.34 seconds (week 8), and 0.22 ± 1.13 seconds (week 12). These results suggest that EEDK has a prolonged and positive effect on tear film rupture time.

**Figure 2.**
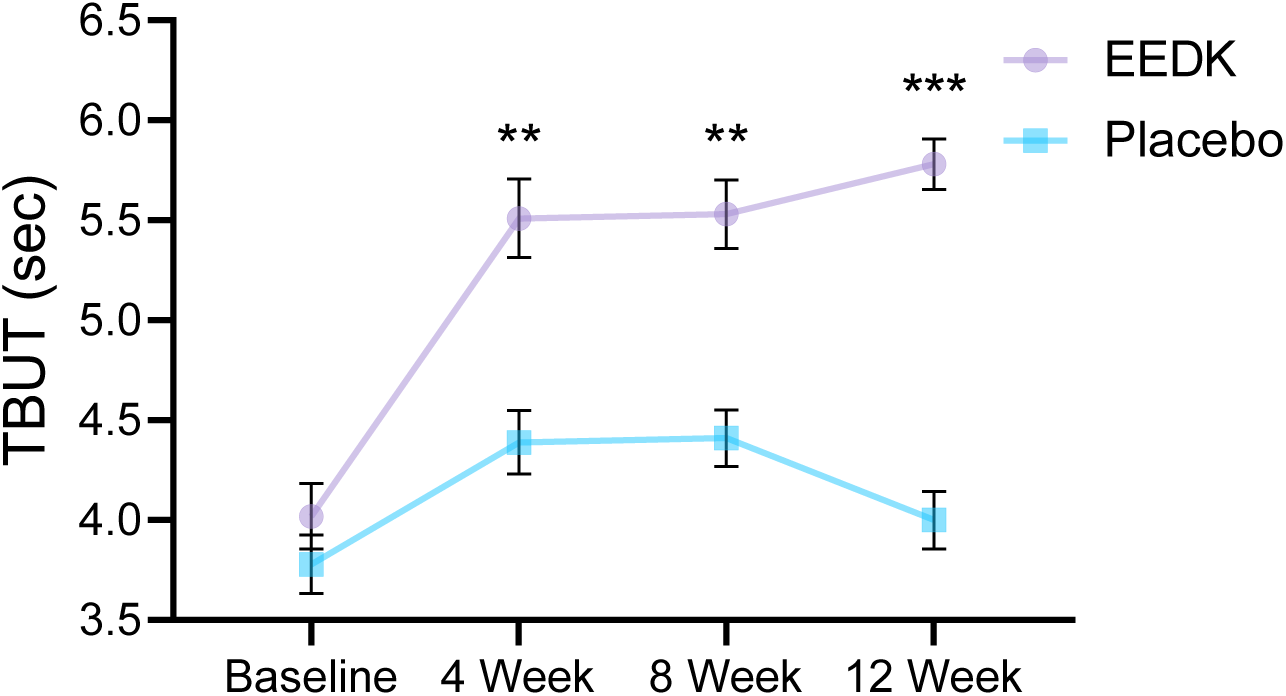
Tear film break-up time (TBUT) values in the EEDK and placebo groups. Significant difference between two groups at p < 0.01 (**); p < 0.001 (***). Values are expressed as mean ± SEM.

**Table 2.** The changes of TBUT, IOP, Schirmer test, and corneal fluorescein stain score in two groups for 12 weeks of treatment.

| Parameters | Baseline | 4 week | 8 week | 12 week | *p*-Value<sup>&</sup> | *p*-Value<sup>\$</sup> |
| --- | --- | --- | --- | --- | --- | --- |
| Visual acuity |  |  |  |  |  |  |
| EEDK group | 0.81 ± 0.16 | 0.81 ± 0.14 | 0.80 ± 0.12 | 0.82 ± 0.12 | 0.2054 | 0.0038<sup>a</sup> |
| Placebo group | 0.75 ± 0.15 | 0.73 ± 0.14 | 0.73 ± 0.12 | 0.73 ± 0.12 |  |  |
| TBUT (s) |  |  |  |  |  |  |
| EEDK group | 4.02 ± 1.08 | 5.51 ± 1.28 | 5.53 ± 1.12 | 5.78 ± 0.83 | <.0001<sup>a</sup> | <.0001 |
| Placebo group | 3.78 ± 0.99 | 4.39 ± 1.08 | 4.41 ± 0.96 | 4.00 ± 0.97 |  |  |
| IOP |  |  |  |  |  |  |
| EEDK group | 21.62 ± 0.43 | 18.16 ± 1.79 | 17.20 ± 1.52 | 16.81 ± 1.09 | <.0001 | <.0001 |
| Placebo group | 22.16 ± 2.03 | 20.88 ± 2.01 | 20.77 ± 2.08 | 20.76 ± 1.68 |  |  |
| Schirmer test (mm) |  |  |  |  |  |  |
| EEDK group | 8.00 ± 3.41 | 9.70 ± 3.52 | 9.49 ± 3.20 | 9.53 ± 3.10 | 0.0009<sup>a</sup> | 0.0287<sup>a</sup> |
| Placebo group | 9.04 ± 3.75 | 8.54 ± 4.13 | 8.33 ± 3.31 | 7.74 ± 3.92 |  |  |
| CS score |  |  |  |  |  |  |
| EEDK group | 3.77 ± 0.92 | 1.77 ± 1.09 | 1.37 ± 0.87 | 1.05 ± 0.49 | <.0001 | <.0001 |
| Placebo group | 3.61 ± 0.77 | 3.07 ± 0.77 | 2.83 ± 0.80 | 2.93 ± 0.65 |  |  |
| NIBUT |  |  |  |  |  |  |
| EEDK group | 4.58 ± 0.71 | 4.72 ± 0.77 | 4.61 ± 0.79 | 4.72 ± 0.82 | 0.5964 | 0.4700 |
| Placebo group | 4.55 ± 0.63 | 4.65 ± 0.65 | 4.63 ± 0.64 | 4.59 ± 0.99 |  |  |
| MGS score |  |  |  |  |  |  |
| EEDK group | 1.56 ± 0.67 | 1.58 ± 0.70 | 1.58 ± 0.70 | 1.56 ± 0.67 | 0.4442 | 0.5359 |
| Placebo group | 1.41 ± 0.62 | 1.43 ± 0.62 | 1.48 ± 0.66 | 1.46 ± 0.66 |  |  |
| OSDI |  |  |  |  |  |  |
| EEDK group | 27.69 ± 22.24 | 16.03 ± 13.60 | 12.32 ± 10.73 | 10.24 ± 11.17 | 0.1072 | 0.3309 |
| Placebo group | 21.91 ± 20.04 | 13.97 ± 15.94 | 12.36 ± 14.52 | 11.17 ± 14.29 |  |  |

<sup>a</sup>: *p*<0.05 between EEDK group and placebo group; <sup>&</sup>: Compared between groups, *p*-value for Wilcoxon rank sum test; <sup>§</sup>: Compared between groups; *p*-value for GLM adjusted baseline,
drinking, smoking, sleeping time, smartphone (computer) usage time, exercise, and central corneal thickness. TBUT: Tear film break-up time; IOP: Intraocular Pressure; CS: Corneal stain; NIBUT: non-invasive tear film break-up time; MGS: Meibomian gland secretion; OSDI: Ocular surface disease index.

The IOP test was performed at weeks 4, 8, and 12 in the EEDK group. Significant differences in IOP improvement were observed between the EEDK and placebo groups (week 4, p < 0.0001; week 8, p < 0.0001; and week 12, p < 0.0001; **Table 2** and **Figure 3**). Compared with the baseline value, the IOP in the EEDK group showed a significant decrease after 4 weeks (-3.45 ± 1.77 mmHg), 8 weeks (-4.42 ± 1.55 mmHg), and 12 weeks (-4.80 ± 1.16 mmHg); a significant downward trend was observed. For the placebo group, IOP slightly decreased compared with the baseline value at week 4 (-1.28 ± 1.34 mmHg), week 8 (-1.39 ± 0.89 mmHg), and at week 12 (-1.40 ± 0.86) (**Figure 3**). In conclusion, the EEDK group demonstrated significant and consistent improvements in intraocular pressure (IOP) compared with the placebo group over the course of 12 weeks, indicating the effectiveness of EEDK in reducing IOP.

**Figure 3.**
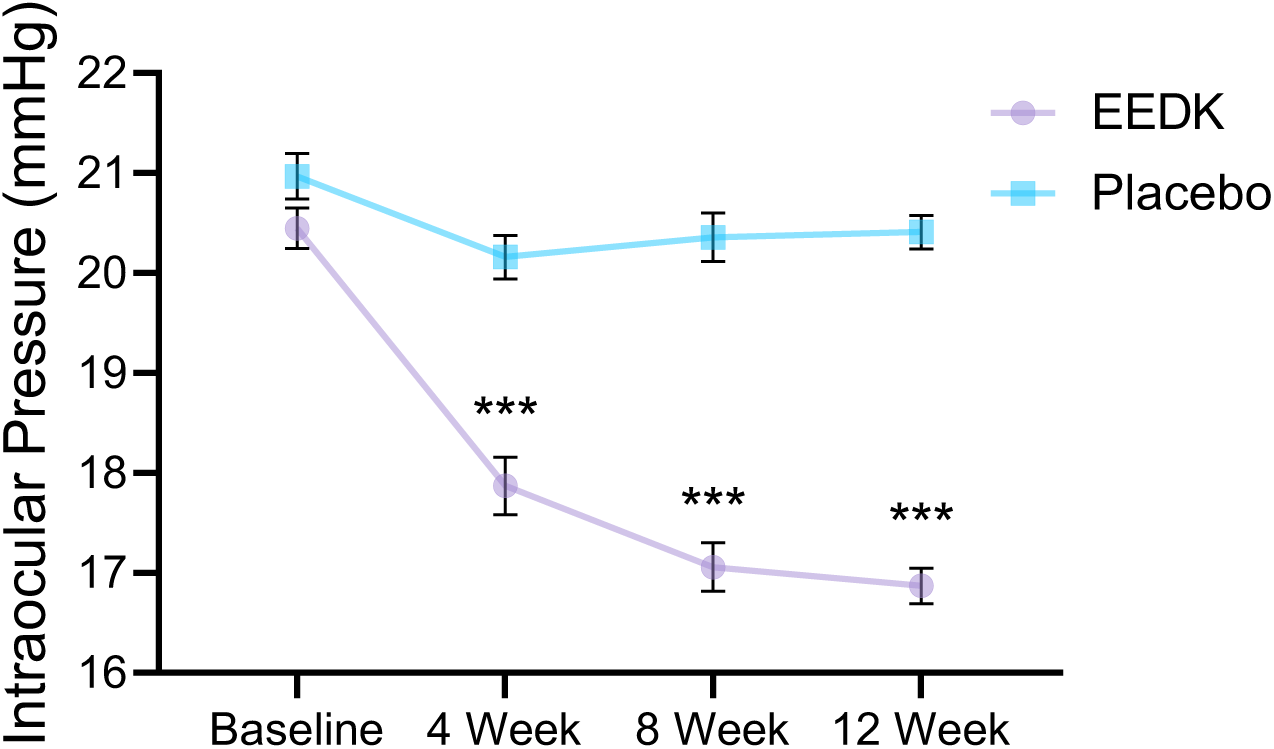
Intraocular pressure (IOP) values in the EEDK and placebo groups. Significant difference between two groups at p < 0.001 (***). Values are expressed as mean ± SEM.

**Figure 4** compares the corneal staining score (CS) at 0, 4, 8, and 12 weeks. After 4 weeks, the EEDK group exhibited a significant decrease of 2.00 ± 1.31 in CS, while the placebo group showed a decrease of 0.54 ± 0.91, indicating a statistically significant difference between the two groups (p < 0.0001). Similarly, after 8 weeks, the EEDK group demonstrated a significant decrease of 2.40 ± 1.22 in CS, compared with a decrease of 0.78 ± 1.15 in the placebo group, revealing a statistically significant difference between the two groups (p < 0.0001). Finally, after 12 weeks, the EEDK group exhibited a significant decrease of 2.72 ± 1.08 in CS, while the placebo group showed a decrease of 0.67 ± 0.94, demonstrating a statistically significant difference between the two groups (p < 0.0001) (**Table 2** and **Figure 4**). Over 12 weeks, EEDK intake led to a statistically significant improvement in the corneal staining score compared with the placebo group, suggesting the efficacy of EEDK in reducing corneal staining.

**Figure 4.**
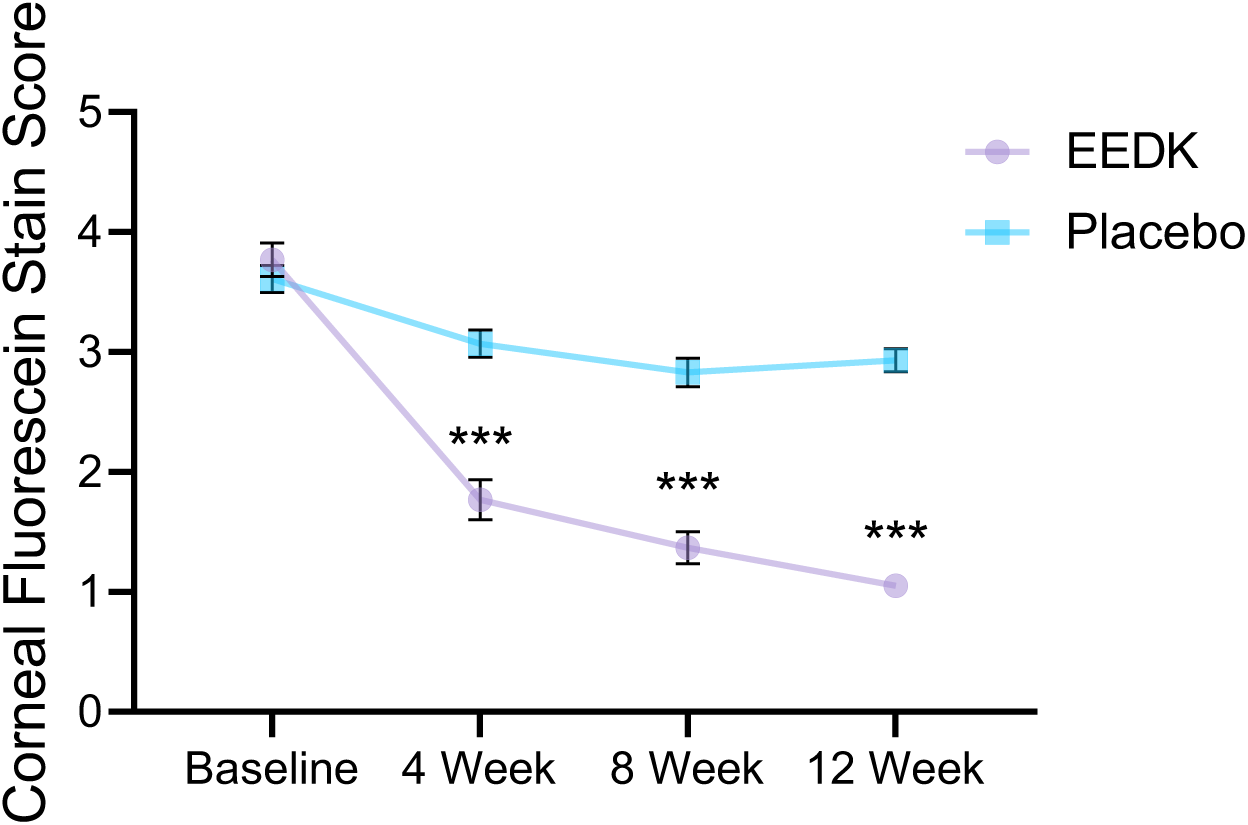
Corneal fluorescein staining scores in the EEDK and placebo groups. Significant difference between two groups at p < 0.001 (***). Values are expressed as mean ± SEM.

The administration of EEDK resulted in a gradual improvement in tear production as assessed by the Schirmer test. In the EEDK group, the baseline scores increased by 1.70 ± 4.51 (week 4), 1.49 ± 3.84 (week 8), and 1.53 ± 4.72 (week 12). Conversely, the placebo group exhibited a slight decrease in Schirmer’s score, with reductions of 0.50 ± 5.19 (week 4), 1.30 ± 4.82 (week 8), and 1.53 ± 4.72 (week 12) (**Table 2**). Significant differences in the Schirmer test value improvement between the EEDK group and placebo group were observed at 4, 8, and 12 weeks (week 4, p = 0.0363; week 8, p = 0.0203; week 12, p = 0.0009; **Table 2** and **Figure 5**). These findings indicate that EEDK intake effectively enhanced the secretion of basal tears, suggesting a positive effect on dry eye.

**Figure 5.**
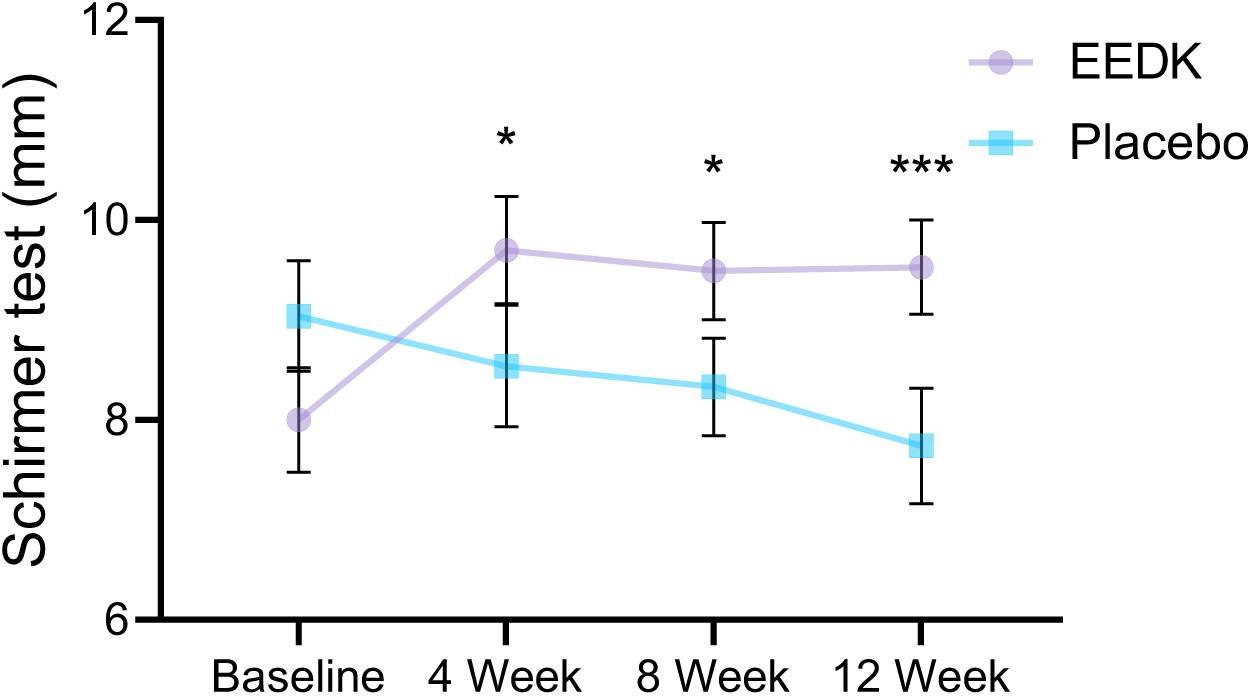
Schirmer test values in the EEDK and placebo groups. Significant difference between two groups at p < 0.05 (*); p < 0.001 (***). Values are expressed as mean ± SEM.

Analysis of changes in NIBUT showed that after 12 weeks of ingestion, the EEDK group experienced an increase of 0.14 ± 1.01 sec, while the placebo group exhibited an increase of 0.03±1.08 sec; however, no statistically significant difference was observed between the two groups (**Table 2**).

Similarly, when analyzing the change in Meiboscore of the eyes after 12 weeks, the EEDK group showed an increase of 0.00 ± 0.31, while the placebo group displayed an increase of 0.04 ± 0.21; however, there was no statistically significant difference observed between the two groups (**Table 2**).

The OSDI values were significantly reduced in both the EEDK and placebo groups after 4, 8, and 12 weeks (**Figure 6**). Both groups demonstrated a consistent downward trend compared with baseline values throughout the study duration (p < 0.001). However, no statistically significant differences were observed between groups (**Table 2**). The EEDK group displayed a more pronounced downward trend than the placebo group. EEDK treatment appears to provide substantial relief and therapeutic benefits for the management of dry eye disease.

**Figure 6.**
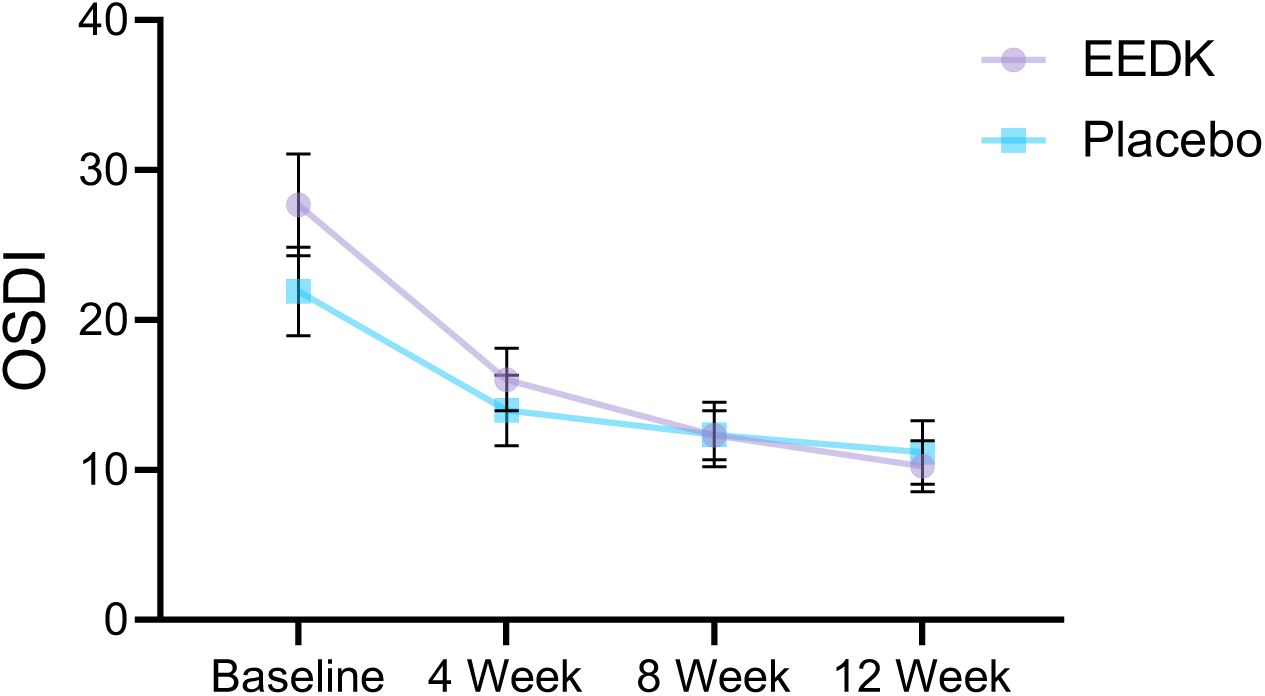
Ocular surface disease index (OSDI) values in the EEDK and placebo groups. Values are expressed as mean ± SEM.

Analysis of changes in visual acuity was measured at 0, 4, 8, and 12 weeks. After 12 weeks of intake, the EEDK group increased by 0.00 ± 0.08 (p = 0.4221), whereas the control group decreased by 0.02 ± 0.10 (p = 0.0852), demonstrating a statistically significant difference between the groups (p = 0.0038; **Table 2**).

The evaluation of improvement was conducted by investigators and trial subjects at weeks 4, 8, and 12. In the analysis of the investigator’s improvement evaluation scores, the EEDK group scored 2.58 ± 0.50 after 4 weeks of intake, while the control group scored 2.87 ± 0.34, indicating a statistically significant difference between the groups (P = 0.0024). Similarly, after 8 weeks of intake, the EEDK group scored 2.05 ± 0.21, and the control group scored 2.65 ± 0.48, with a statistically significant difference between the groups (P < 0.0001). After 12 weeks of intake, the EEDK group scored 2.02 ± 0.15, and the control group scored 2.70 ± 0.47, demonstrating a statistically significant difference between the groups (P < 0.0001) (**Table 3** and **Figure 7**). However, the analysis of trial subjects’ self-reported improvement scores did not reveal any statistically significant differences between the EEDK and control groups after 4, 8, and 12 weeks of intake.

**Figure 7.**
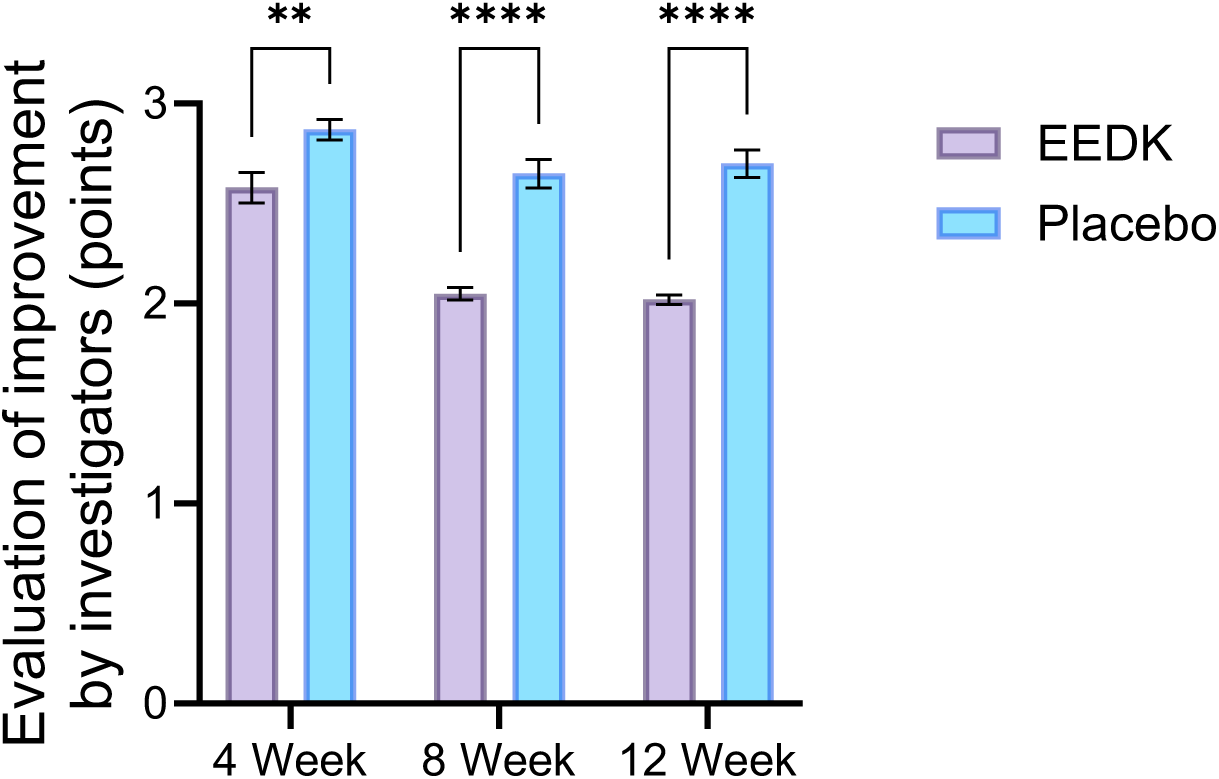
Evaluation of improvement by investigators in the EEDK and placebo groups. Values are expressed as mean ± SEM.

**Table 3.**
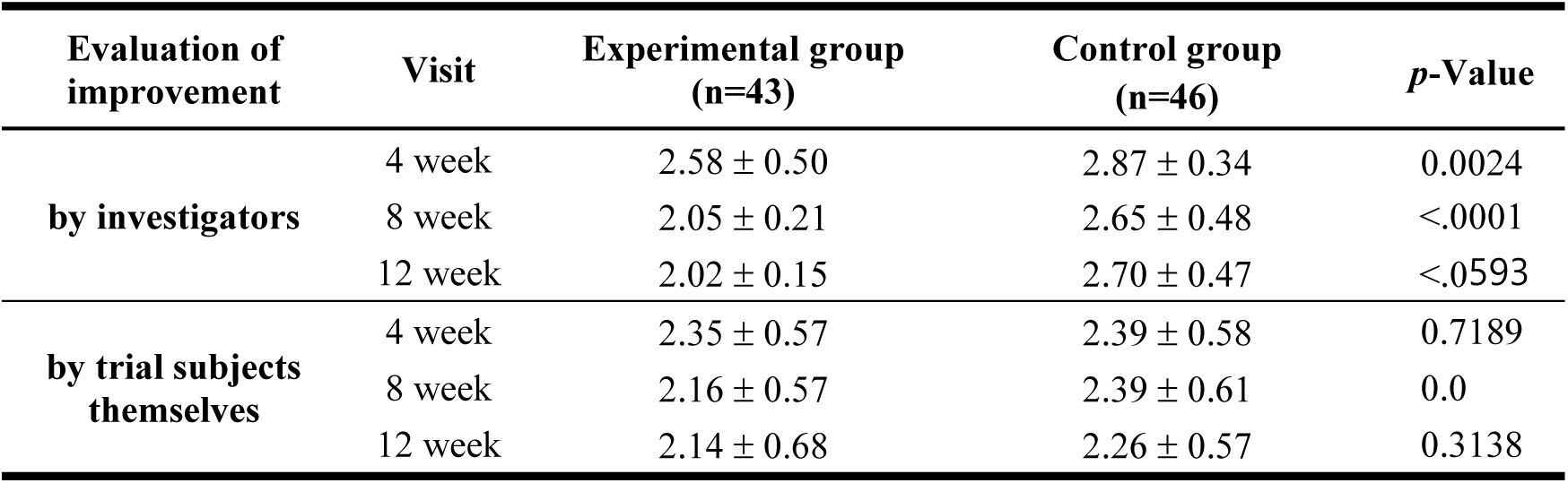
Evaluation of improvement by investigators and trial subjects at each visit.

| Evaluation of improvement | Visit | Experimental group (n=43) | Control group (n=46) | *p*-Value |
| --- | --- | --- | --- | --- |
| by investigators | 4 week | 2.58 ± 0.50 | 2.87 ± 0.34 | 0.0024 |
|  | 8 week | 2.05 ± 0.21 | 2.65 ± 0.48 | <.0001 |
|  | 12 week | 2.02 ± 0.15 | 2.70 ± 0.47 | <.0001 |
| by trial subjects themselves | 4 week | 2.35 ± 0.57 | 2.39 ± 0.58 | 0.7189 |
|  | 8 week | 2.16 ± 0.57 | 2.39 ± 0.61 | 0.0633 |
|  | 12 week | 2.14 ± 0.68 | 2.26 ± 0.57 | 0.3138 |

### 3.3. Safety evaluation

The safety analysis included 100 patients (50 in the EEDK group and 50 in the control group) who received at least one dose of the investigational product. Throughout the trial period, two adverse events (AEs) were reported in two patients receiving EEDK (4%), while nine AEs were observed in three patients receiving the placebo (6%); however, no significant differences were found between the two groups (p=1.0000) (**Table 4**). In the experimental group, the reported adverse reactions included infections and parasitic infections (2.00%), and respiratory, thoracic, and mediastinal disorders (2.00%). The control group experienced nine adverse reactions, primarily gastrointestinal disorders (4.00%) and bacterial and parasitic infections (4.00%). No serious adverse events (SAEs) were recorded, and no participants dropped out due to adverse reactions. No clinically significant changes were observed in the vital signs of either group, and the reported changes in blood chemistry (**Table 5**) and urine test results (**Table 6**) were not considered clinically significant between the two groups. Regarding visual acuity, anterior segment examination, and intraocular pressure, neither group demonstrated any clinically significant changes from the baseline.

**Table 4.** Incidence and classification of adverse events in the safety set.

|  | **Experimental group (n=50)** |  | **Control group (n=50)** |  | **Sum (n=100)** |  | ***p*-Value** |
| --- | --- | --- | --- | --- | --- | --- | --- |
|  | n (%) | Occurrences | n (%) | Occurrences | n (%) | Occurrences |  |
| Adverse event (AE) | 2 (4.00) | 2 | 3 (6.00) | 9 | 5 (5.00) | 11 | 1.0000 |
| Serious adverse event (SAE) | 0 (0.00) | 0 | 0 (0.00) | 0 | 0 (0.00) | 0 | - |
| Dropout due to adverse reaction | 0 (0.00) | 0 | 0 (0.00) | 0 | 0 (0.00) | 0 | - |

**Table 5.** Highlights of blood test index before and after treatment.

|  |  | Experimental group | Control group | *p*-Value |
| --- | --- | --- | --- | --- |
| AST(GOT)<br>(IU/L) | *Week 0* | 24.30±6.15 | 27.60±9.27 | 0.0997 |
|  | *Week 12* | 24.38±6.02 | 27.79±9.65 | 0.9851 |
| ALT(GPT)<br>(IU/L) | *Week 0* | 20.58±11.96 | 24.56±12.24 | 0.0577 |
|  | *Week 12* | 20.11±10.54 | 25.42±14.65 | 0.5708 |
| Total protein<br>(g/dL) | *Week 0* | 7.39±0.36 | 7.44±0.32 | 0.2760 |
|  | *Week 12* | 7.34±0.40 | 7.43±0.29 | 0.4879 |
| Albumin<br>(g/dL) | *Week 0* | 4.34±0.23 | 4.37±0.21 | 0.5256 |
|  | *Week 12* | 4.25±0.24 | 4.28±0.28 | 0.8689 |
| Glucose<br>(mg/dL) | *Week 0* | 98.52±10.77 | 98.94±10.70 | 0.5210 |
|  | *Week 12* | 99.94±12.01 | 101.83±13.28 | 0.7741 |
| ALP (IU/L) | *Week 0* | 70.82±18.72 | 72.42±16.45 | 0.6509 |
|  | *Week 12* | 67.13±18.99 | 71.27±17.28 | 0.3397 |
| CPK (IU/L) | *Week 0* | 106.38±58.38 | 123.40±94.01 | 0.3945 |
|  | *Week 12* | 109.94±60.86 | 123.02±90.07 | 0.4149 |
| Creatinine<br>(mg/dL) | *Week 0* | 0.70±0.15 | 0.71±0.15 | 0.6607 |
|  | *Week 12* | 0.71±0.15 | 0.72±0.16 | 0.5267 |
| Uric acid<br>(mg/dL) | *Week 0* | 5.04±1.32 | 4.99±1.41 | 0.9780 |
|  | *Week 12* | 4.95±1.17 | 4.97±1.51 | 0.8231 |

**Table 6.**
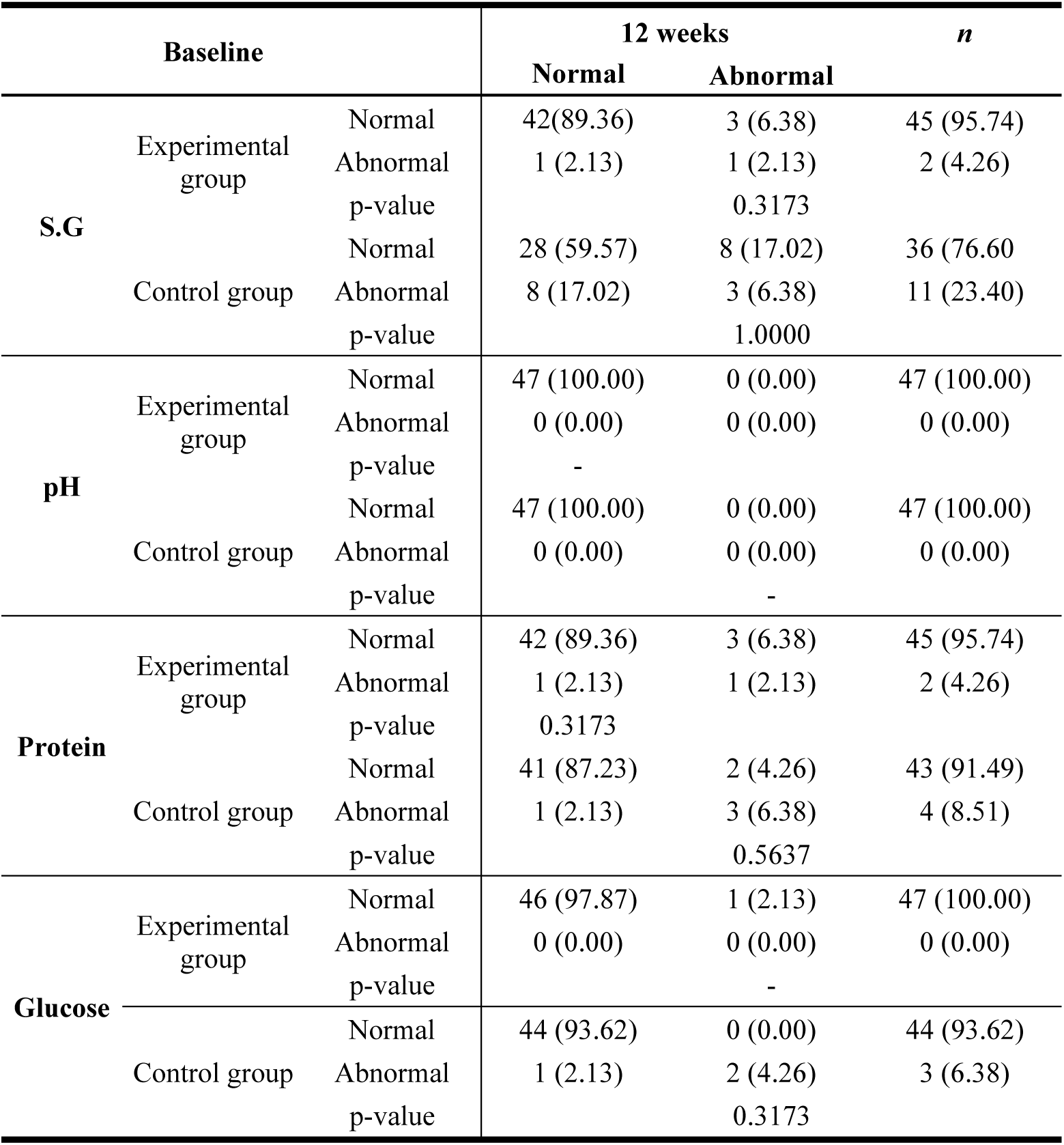
Summary of the patients’ urine test results.

## 4. Discussion

The recent increase in the incidence of DED can be attributed to an increase in poor eye care habits and inadequate knowledge of eye hygiene [21]. DED is a group of syndromes that manifests as dry eyes, photophobia, and excessive tear secretion [22]. Multiple kinetic abnormalities in tear texture, decreased tear secretion, and compromised tear film stability are pathological substrates of DED. These abnormalities can persistently disrupt the repair and defense mechanisms of the ocular surface, and inhibit sensory nerves [23]. Consequently, patients with prolonged DED with various symptoms and recurrent episodes frequently experience a significant decline in their quality of life [24].

Currently, artificial tears are the primary therapeutic intervention for DED, particularly in postoperative patients. These eye drops are intended to alleviate symptoms by increasing ocular surface moisture and lubrication, thereby enhancing tear film stability [25]. It is essential to note, however, that artificial tears provide only transient relief and lack inherent tissue-repair or anti-inflammatory effects, thereby limiting their overall effectiveness [26]. In addition, the prolonged use of artificial tears may result in diminished therapeutic benefits and an increased risk of adverse drug reactions, ultimately impeding the improvement of ocular symptoms in patients [27]. Therefore, alternative therapeutic options for DED that do not cause any local side effects are clearly needed. Such treatment methods should target the underlying pathogenesis of DED and provide long-term benefits to patients. Future research efforts should focus on developing novel interventions that not only alleviate ocular discomfort, but also promote tissue repair, reduce inflammation, and enhance overall ocular health [28]. By addressing these critical gaps, we hope to significantly improve the management and outcomes of DED patients, thereby enhancing their quality of life.

The traditional medicinal uses of *D. kaki* leaves are well documented because of their potent radical-scavenging, antioxidant, and immune-modulatory properties [13–15]. These leaves contain a rich assortment of bioactive constituents, including various triterpenoids, flavonoids, such as kaempferol and quercetin derivatives, and other compounds [29–31]. Considering the involvement of free radicals, radiation, and fatigue in numerous ocular diseases [32], *D. kaki* leaves have the potential to prevent and treat these conditions. In our previous studies, we highlighted the positive effects of EEDK on clinical indicators of dryness. Specifically, EEDK has demonstrated efficacy in mitigating corneal neovascularization induced by alkali burns in vivo and ameliorating retinal degeneration both in vitro and in vivo [33, 34]. Additionally, EEDK has shown promise in enhancing clinical indicators of dryness in a mouse model of dry eye induced by benzalkonium chloride. Administration of EEDK resulted in longer tear breakup time, reduced corneal fluorescein staining scores, increased tear volume, and smoother epithelial cells [35]. Notably, EEDK treatment has also shown potential benefits in preventing dry eye disease by increasing goblet cell density and exerting anti-inflammatory effects in a mouse model [36]. The success of these animal experiments has paved the way for future clinical trials investigating the therapeutic potential of EEDK in humans.

In light of these encouraging preclinical findings, we conducted a new prospective clinical trial of EEDK to test the hypothesis that administering the EEDK to patients with DED would result in a greater improvement in their dry eye symptoms than the placebo group.

The TBUT and Schirmer’s test are commonly used to evaluate tear film stability and tear production, respectively. Assessment of TBUT and measurement of tear secretion using the Schirmer’s test are widely used clinical indices to evaluate the condition of the ocular surface. Previous studies conducted in mouse models of dry eye have demonstrated that EEDK improves both TBUT and Schirmer’s test scores [35, 36]. Consistent with these findings, our study observed a similar pattern in the efficacy of EEDK as observed in previous animal experiments. Significant improvements in tear film stability and tear production were evident 4 weeks after initiating EEDK treatment, as indicated by the improvement in TBUT and Schirmer’s test results. Importantly, EEDK exhibited superior outcomes in terms of TBUT and Schirmer’s test scores compared with the placebo group, suggesting its efficacy as an effective treatment for DED. Overall, our study provides further evidence supporting the conclusion that EEDK intake can enhance tear film stability and production in individuals diagnosed with DED.

Ocular surface health is commonly evaluated using fluorescein staining, a well-established method. Fluorescein is unable to penetrate healthy epithelial lipid layers, but readily stains damaged corneal and conjunctival surfaces with disrupted cell-to-cell junctions [37]. Thus, corneal fluorescein staining serves as an indicator of ocular surface inflammation and has been shown to improve with anti-inflammatory therapies. In our study, the administration of EEDK for 12 weeks significantly improved corneal fluorescein staining compared with the placebo group. It is noteworthy that the leaves of *D. kaki* contain an abundance of phenolic compounds, which suppress the production of inflammatory mediators, such as nitric oxide and prostaglandin E2, as well as proinflammatory cytokines like TNF-a and IL-1b, in macrophages [29, 38]. Based on the findings of these studies, we suggest that the anti-inflammatory properties of EEDK may significantly reduce tear film cytokine concentrations, thereby leading to a reduction in corneal staining scores, which is indicative of diminished ocular surface inflammation.

Several studies have documented an association between elevated IOP and DED [31]. It has been proposed that factors linked to DED, such as inflammation and ocular surface damage, may contribute to increased IOP [31]. Furthermore, some individuals with DED exhibit elevated IOP. Our previous study demonstrated the potential of EEDK to reduce IOP in a microbead-induced ocular hypertensive glaucoma model and in a DBA/2 mouse model that developed an age-dependent glaucomatous phenotype [17]. In the current study, a significant decrease in IOP was observed in the EEDK group compared with the placebo group between weeks 4 and 12. Individuals with DED and elevated IOP may benefit from treatment approaches that target both conditions simultaneously. If a therapeutic drug can effectively lower IOP while improving tear film stability and prolonging tear film rupture, it could provide valuable benefits for patients with DED. Although the mechanism remains unknown, our findings show that EEDK has the potential to be a highly effective treatment for DED by simultaneously improving tear film stability and lowering IOP.

In this study, we observed a decrease in OSDI scores in both the EEDK and control groups. The decrease in the OSDI and improvement in the DED index, specifically in the EEDK group, can be attributed to the consumption of the drug of interest. However, the decrease in OSDI in the control group is difficult to explain and was likely influenced by the placebo effect. The characteristics of the questionnaire used in the survey suggested that the placebo effect played a significant role in the observed results [39]. It is highly probable that participants in the control group experienced improvement based on their belief in receiving beneficial treatment, despite not receiving active intervention.

Our findings suggest that EEDK holds great promise as a highly effective and safe natural plant component with high concentrations of free radical scavengers and anti-inflammatory mediators. By reducing the damaging effects of free radicals and radiation on the ocular surface, EEDK has the potential to alleviate dry eye symptoms, prolong tear film rupture time, decrease IOP, and enhance tear film stability. Moreover, the wide availability of EEDK facilitated the creation and application of these components.

## 5. Conclusion

In conclusion, oral administration of EEDK capsules in the treatment of patients with dry eye has shown remarkable improvements in both objective parameters and subjective symptoms while maintaining a high level of safety. These findings strongly support the wider adoption and promotion of EEDK as an effective and reliable treatment option for individuals suffering from dry eye disease.

## Data Availability

All data produced in the present work are contained in the manuscript

## Author Contributions

**Hyung Bin Hwang**: Investigation, Methodology, Data curation. **Wook-Bin Lee**: Investigation, Writing – original draft, Writing – review & editing. **Kyung-A Kim**: Investigation. **Hong Ryul Ahn**: Investigation. **Tae Kyeom Kang**: Investigation. **Jong Jin Lee**: Methodology, Data curation. **Min Kyoung Kim**: Methodology. **Kui Dong Kang**: Investigation, Conceptualization, Supervision. **Sang Hoon Jung**: Investigation, Conceptualization, Supervision

## Acknowledgements

This work was supported by an intramural grant (2E33531) from the Korea Institute of Science and Technology (KIST), Republic of Korea. Funding is also provided by Whanin Pharm Co., Ltd., Seoul, Korea. The funding organization participated in the study design and conduction, data analysis and interpretation, and preparation of the manuscript.

## Abbreviations

CS: corneal staining
DED: dry eye disease
EEDK: ethanol extract of Diospyros kaki
IOP: intraocular pressure
MGS: meibomian gland secretion
NIBUT: noninvasive tear film breakup time
OSDI: ocular surface disease index
TBUT: tear breakup time

